# PTPRG activates m6A methyltransferase VIRMA to block mitochondrial autophagy mediated neuronal death in Alzheimer’s disease

**DOI:** 10.1101/2022.03.11.22272061

**Authors:** Jiefeng Luo, Xiaohua Huang, Rongjie Li, Jieqiong Xie, Liechun Chen, Chun Zou, Zifei Pei, Yingwei Mao, Donghua Zou

**Affiliations:** Department of Neurology, The Second Affiliated Hospital of Guangxi Medical University, Nanning, Guangxi 530007, People’s Republic of China; Department of Neurology, The Affiliated Hospital of Youjiang Medical University for Nationalities, Baise, Guangxi 533000, People’s Republic of China; Department of Neurology, The Fifth Affiliated Hospital of Guangxi Medical University, Nanning, Guangxi 530022, People’s Republic of China; Department of Biology, Pennsylvania State University, University Park, PA, 16802, USA

**Keywords:** neuronal death ecotone, single-cell technology, spatial transcriptomics, mitochondrial damage depletion, m6A methylation modification, energy and nutrient metabolism imbalance

## Abstract

In Alzheimer’s disease (AD), neuronal death is one of the key pathology. However, the initiation of neuronal death in AD is far from clear, and new targets are urgently needed to develop effective therapeutic methods. This study analyzed sequencing data from single-cell RNAseq and spatialomics, and revealed the impact of global single-cell mapping and cell spatial distribution relationships in early stage of AD. We found that microglia subpopulation Mic_PTPRG can anchor neurons based on ligand-receptor interaction pairs and cause ectopic expression of PTPRG in neurons during AD progression. PTPRG in neurons can bind and upregulate VIRMA expression, which in turn increases the level of m6A methylation, enhances PRKN transcript degradation and represses translation. Repressed PRKN expression blocks the clearance of damaged mitochondria in neurons, which in turn reprograms neuronal energy and nutrient metabolic pathways and leads to neuronal death during AD progression. This study elucidates novel mechanisms, by which the PTPRG-dependent microglia-synaptic modification may play a role in AD, providing a new scientific basis for potential therapeutic targets for AD.

## Introduction

Alzheimer’s disease (AD), the main form of dementia in the elderly, is a progressive and irreversible neurodegenerative disease accompanied by the degeneration and neuronal death (1). The symptoms of AD usually begin with mild memory difficulties and gradually progress to cognitive impairment, language impairment and personality changes as the disease deteriorates (2). The long duration of the disease and the high rate of disability make the patients lose their ability to live independently in the late stages and require complete care from others, which brings a heavy burden to the society and families. In the last decades, studies have found that biological (e.g., aging, gender, weight, etc.), environmental (e.g., lifestyle, toxins, brain injury, etc.) and genetic (e.g., APP, PS1 and PS2 gene mutations in familial AD and susceptibility genetic polymorphisms in disseminated cases) factors play an important role in the pathogenesis of AD (3). Although these findings have greatly improved our understanding of AD, the underlying mechanisms of AD pathogenesis remain elusive, and current treatments can only target symptoms but not alter the disease process.

It is now generally accepted that extracellular aggregation of amyloid-β (Aβ) plaques plays an important role in the pathogenesis of AD (4). In addition, intracellular neurofibrillary tangles (NFT), which consist of hyperphosphorylated TAU proteins, also play an important role in the pathogenesis of AD and are also the most prominent pathological process in neurons (5, 6). In the brain, neurons express a large number of molecules that protect them from inflammatory attacks and induce neurological dysfunction. However, neuronal death is present to various degrees in almost all AD patients (7), suggesting that the ecological niche of neuronal death - both excitatory and inhibitory neurons - may have different degrees of cell death. Notably, neurons are high-energy cells that depend primarily on mitochondria for ATP production and Ca^2+^ homeostasis to maintain action potential (8), whereas mitochondria are highly dynamic organelles that regulate their morphology and distribution through constant division and fusion events (9, 10). Through this process, mitochondria respond to high energy demands and promote neuroprotective effects by eliminating defective mitochondrial components and preventing damage from reactive oxygen species during aging (11-13). However, mitochondrial dysfunction is commonly found in AD (14). Therefore, it is crucial to explore in depth the relationship between neuronal death and mitochondrial damage, which not only helps to further elucidate the pathogenesis of AD, but also may provide potential novel therapeutic targets for the treatment of AD.

However, the human brain is an extremely complex tissue and ordinary research tools are no longer sufficient for our research needs. The recent popularity of single-cell technology and spatial histology has provided us with advanced technological conditions. Single-cell technology allows us to understand and explore diseases with a high-resolution perspective. Spatialomics is another research technology other than single-cell sequencing, which can reveal the influence of cell spatial distribution relationship on diseases by studying the relative position relationship of cells in tissue samples. Thus spatialomics compensates the shortcoming that single-cell sequencing technology cannot obtain information of cell spatial distribution. Currently, single-cell technology and spatial histology have been widely used in the study of brain development and AD. Ugomma C Eze et al. used single-cell techniques to characterize the major cell types of human brain development and the subpopulations of progenitor cells that form the basis of the human cerebral cortex (15). Sabina Kanton et al. used single-cell techniques to reveal dynamic gene regulation features of human brain development (16). Shun-Fat Lau et al. based on single-nucleus transcriptome analysis revealed dysregulation of AD vascular endothelial cells and neuroprotective glial cells (17). In addition, Jos é Fernández Navarro et al. revealed genes associated with mitochondrial dysfunction and stress signaling in AD based on spatial transcriptomics (18).

Further exploration of the ecological niche of neuronal death in AD can help us understand how AD occurs and progresses, and eventually guide the development of new diagnostic and therapeutic strategies. To this end, this study analyzed single-cell RNA sequencing (scRNA-seq) data from AD brains as well as spatial transcriptomics data from AD mice. In contrast to previous studies, we provide new insights and molecular evidence for cellular communication at the single-cell transcriptome level combined with spatial transcriptomics in AD. In particular, microglia and neurons play a dominant role in the intercellular communication network in AD, revealing the development of AD by exploring the relationship between neuronal death and mitochondrial damage.

## Materials and Methods

### Data source

To study transcriptomic changes in various cell types during AD, we collected the following AD single-cell RNA sequencing (scRNAseq) datasets from the Gene Expression Omnibus (www.ncbi.nlm.nih.gov/geo) database: GSE138852 on the GPL18573 platform, GSE146639 on the GPL21697 platform, GSE147528, GSE157827, GSE163577, GSE174367 and GSE181279 on the GPL24676 platform, GSE160936 on the GPL20301 platform. In addition, there is a set of data from Hansruedi Mathys’ group of professors (19). Samples for all data included tissue samples from prefrontal cortex, internal olfactory cortex, superior parietal lobe, superior frontal gyrus, caudal internal olfactory cortex, somatosensory cortex, hippocampus, superior frontal cortex, and peripheral blood mononuclear cells, involving a total of 85 AD samples and 83 control samples. In addition, spatial transcriptome data were obtained from GSE152506 on GPL19057 platform for mouse coronal slices including 3, 6, 12 and 18 month old AppNL-G-F KI mice (experimental group: n=6) and C57Bl/6J mice (control group: n=6).

### Cell clusters and differentially expressed genes

The Seurat package (20) was used for cell clustering of scRNA-seq data and to visualize clusters using the UMAP package (21). To define marker genes for clusters, clusters featuring similar marker genes were manually combined into one cell type, followed by the use of gene markers for known cell types to generate initial partitions of cells into broad cell clusters. Using the “FindAllMarkers” function, the differentially expressed mitochondrial genes in each cell type were identified. Differences associated with P<0.05 were considered significant.

### Mapping of spatial transcriptomics metadata

The spatial transcriptomics (ST) technique places histological tissue sections on ST slides, which consist of 1007 ST spots covering the slides. In addition, clustered ST spots were plotted onto hematoxylin and eosin (H&E) stained images with specific colors representing the respective clusters. We performed clustering by the UMAP method based on the morphology observed in the H&E-stained histological tissue sections.

### Identification of Cluster-Specific Genes in Spatial Transcriptomics

For each identified cluster, differentially expressed genes (DEGs) associated with all other spots were identified. A list of spatially clustered genes was first generated for all genes differentially expressed in the ST clusters (logFC > 0.25, adjusted P-value < 0.05).

### Single-cell trajectory analysis

The R package Monocle 3 was applied to align cells along the trajectory in pseudo-time (https://cole-trapnell-lab.github.io/monocle3). After clustering the cells using the above method, the dimensionality was reduced and the results were visualized using the UMAP method. Subsequently, cells were sorted according to their progression through the developmental program. monocle measured this progression in pseudo-time. In this study, single-cell trajectory analysis of cell subtypes was performed as needed.

### Functional enrichment analysis of cell clusters

Gene Ontology (GO) enrichment of cell clusters and Kyoto Encyclopedia of Genes and Genomes (KEGG) pathway analysis were used to determine the biological significance of each cell type. GO and KEGG pathway analysis was performed for marker genes for each cell subpopulation using the clusterProfiler package (22). Results associated with P<0.05 were considered significant.

### Analysis of intercellular communication

Intercellular communication analysis was performed using iTALK in the R package. Receptor-ligand interactions were determined using the STRING database of protein-protein interactions (23).

### Molecular docking of ligands and receptors

To clarify the target binding ability between ligand-receptor axes, molecular docking studies were performed using Hex 8.0.0 software (24). Docking models were visualized using Pymol software (25).

## Results

### Global single-cell and spatial landscapes of AD and healthy brains

The analytical flow of this study is shown in Figure 1A. We constructed a large-scale global single-cell landscape on AD brain tissue and healthy brain tissue. By cell clustering analysis, a total of 252 cell clusters were obtained and identified into 19 cell types based on known markers, including excitatory neurons (ExNeu), oligodendrocytes (Oli), inhibitory neurons (InNeu), microglia (Mic), pyramidal cells (PyrNeu), oligodendrocyte precursor cells (Opc), CD8^+^T cells (CD8.T), astrocytes (Ast), endothelial cells (En), CD4^+^T cells (CD4.T), macrophages (Mac), pericytes (Per), Schwann cells (Sch), Purkinje neurons (PurkinjeNeu), initial T cells (Naive.T), ventricular meningeal cells (Ependymal), fibroblast-like cells (Fib_ like), smooth muscle cells (SMC), and B cells (B) (Figure1 B). Each cell type showed specific expression of markers for known cell types (Figure 1C). Subsequently, differential expression analysis was used to further explore the differentially expressed genes for each cell type in AD. Notably, there was significant dysregulation of mitochondrial gene expression in both excitatory and inhibitory neurons in AD compared to controls (Figure 1D), and there may be neuronal mitochondrial damage depletion-mediated cell death. Further by spatial transcriptome techniques, we could see a significant dysregulation of mitochondrial gene expression in AD relative to controls (Figure 1 E, F).

**Figure 1.**
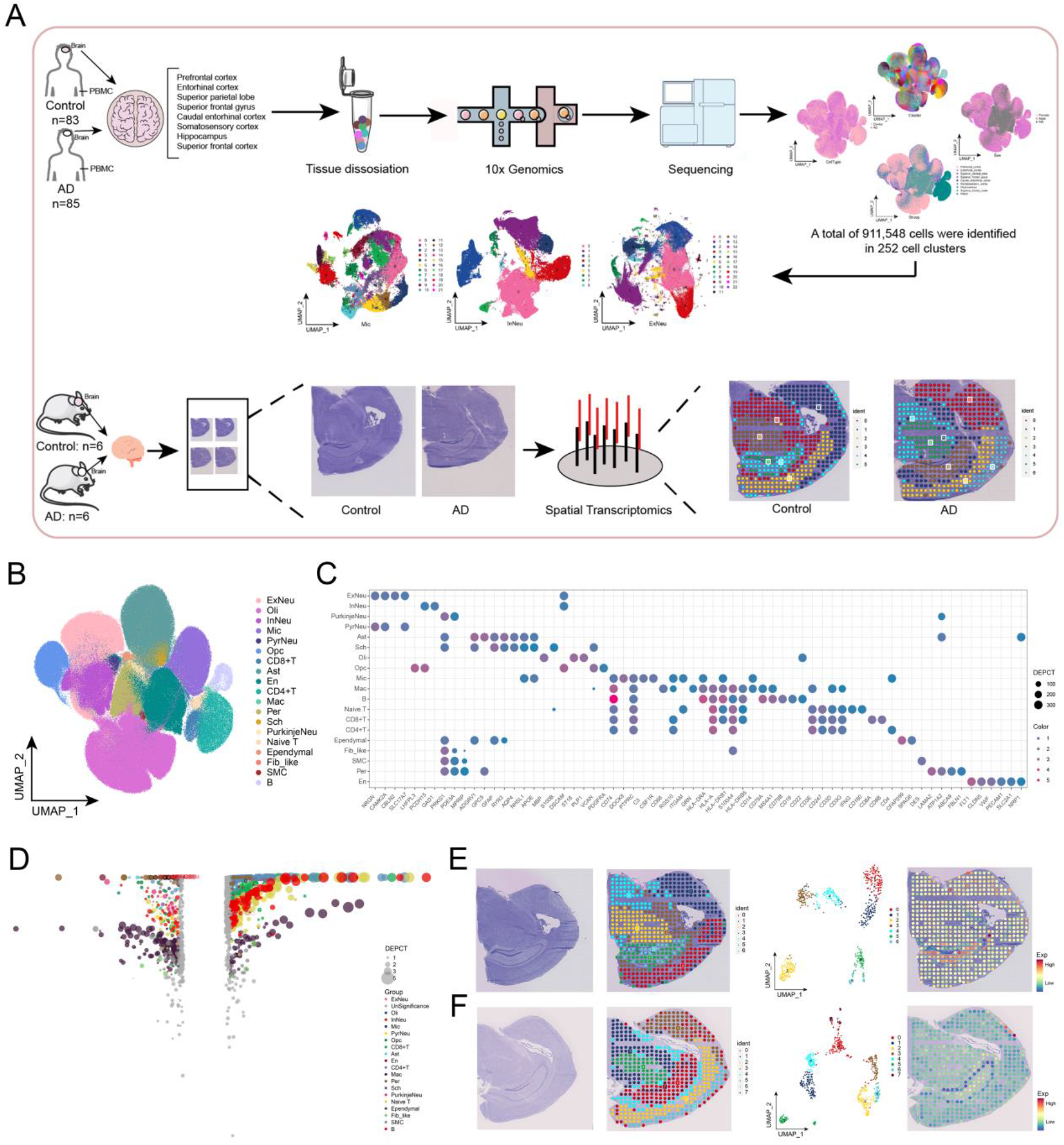
Single-cell transcriptome and spatial transcriptome construction of global single-cell and spatial landscapes of AD and healthy brains. A. Flow chart of this study. B. Single-cell atlas mapping cell types. C. Vesicle map showing global brain tissue single-cell each cell type corresponding to CellMarker. D. Multi-group volcano map showing expression differences of each cell type in AD relative to healthy donor brain cells. E. AD mouse model section HE staining-ST mapping-ST UMAP-Mitochondrial gene expression. F. Healthy mouse model section HE staining-ST mapping-ST UMAP-Mitochondrial gene expression.

### ExNeu_PRKN_VIRMA subpopulation progressively increases in abundance and reaches the highest percentage of excitatory neurons in AD progression

Mitochondria are organelles essential for synaptic function, and their dysfunction is one of the early pathological features of AD. Moreover, there is growing evidence that mitochondrial defect is a key factor contributing to synaptic failure and degeneration in AD. Therefore, we extracted cells with dysregulated mitochondrial gene expression (excitatory and inhibitory neurons) for re-clustering. By analysis, a total of 16 subgroups of excitatory neuronal cells were obtained. Among them, the ExNeu_PRKN_VIRMA subpopulation had the highest percentage in AD patients (Figure 2A). Therefore, we explored the expression as well as co-expression of parkin RBR E3 ubiquitin protein ligase (PRKN) and VIRMA in the single cell profiles of ExNeu_PRKN_VIRMA subpopulation (Figure 2B). In addition, spatial transcriptome profiles of the ExNeu_PRKN_VIRMA subpopulation showed the expression of marker genes of excitatory neurons (*NRGN*), *PRKN* and *VIRMA* in control and AD, respectively. Compared to control, *NRGN, PRKN* and *VIRMA* possessed higher and more extensive expression in AD (Figure 2C). Notably, the abundance of ExNeu_PRKN_VIRMA subpopulation gradually increased with the progression of AD (Figure 2D). Furthermore, the proposed chronological values of excitatory neuron progression from Control to AD were mapped in the single-cell atlas by the proposed chronological analysis (Figure 2E), and the developmental trajectory of excitatory neurons was shown (Figure 2F), and it could be observed that the ExNeu_PRKN_VIRMA subpopulation was at the end of excitatory neuron development. Enrichment analysis showed that the dysregulated genes of ExNeu_PRKN_VIRMA subpopulation were significantly involved in BP such as mitochondrial to lysosomal translocation, mitochondrial autophagy and pathways such as axon guidance, glutamatergic synapses, cell adhesion molecules, calcium signaling pathway, Wnt signaling pathway and Rap1 signaling pathway (Figure 2G). We can see that mitochondrial autophagy of ExNeu_PRKN_VIRMA is inhibited in the pathway map (Figure 2H).

**Figure 2.**
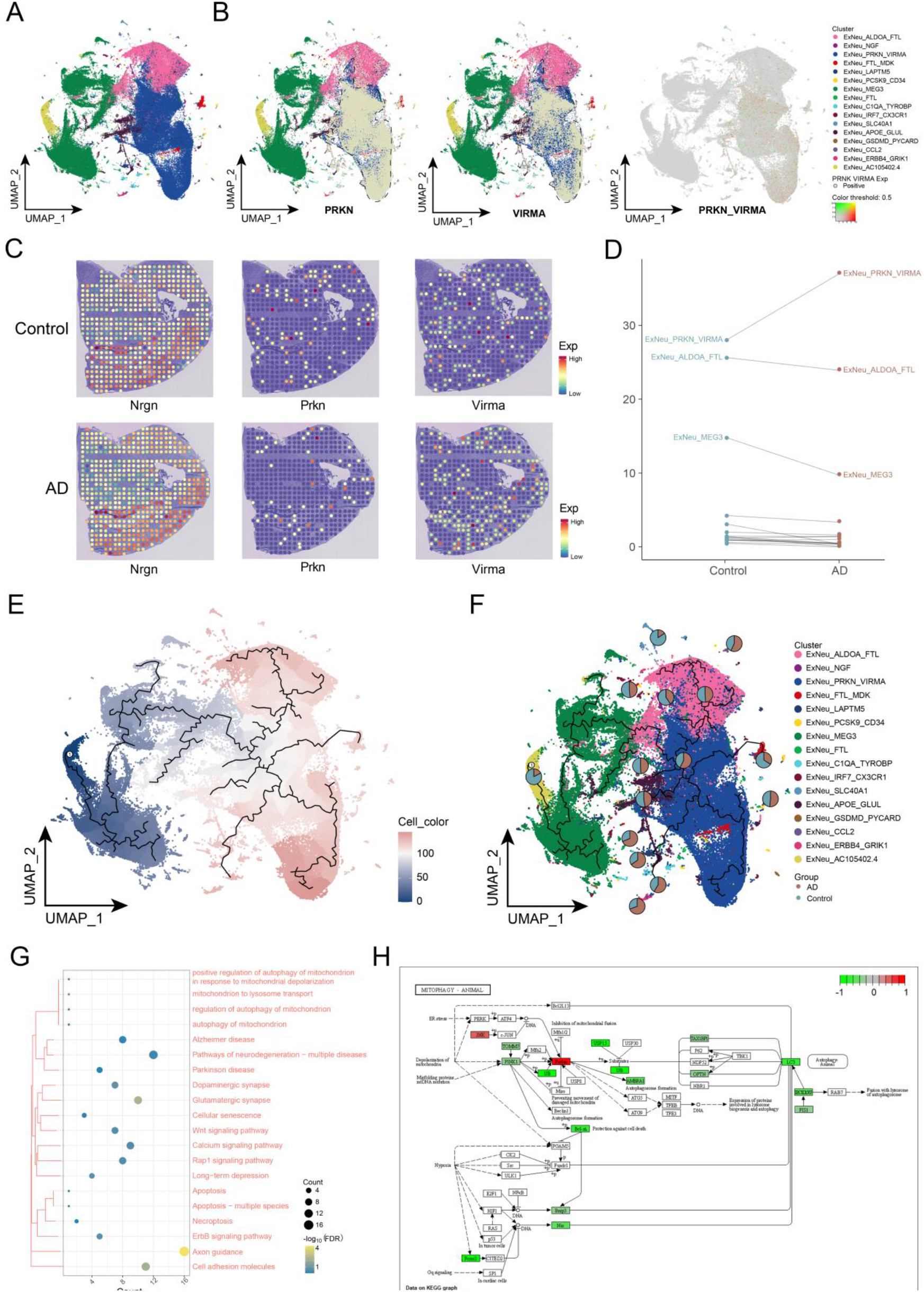
Role of excitatory neuron subpopulations in AD development. A. Single-cell atlas mapping excitatory neuron subpopulations. B. Single-cell atlas demonstrating PRKN and VIRMA expression and co-expression. C. ST atlas demonstrating expression of marker genes *NRGN* and *PRKN* and *VIRMA* in excitatory neuron subpopulations. D. Dotted-line plots demonstrating excitatory neuron subpopulations in AD-Control. E. Single-cell atlas mapping the proposed chronological values of excitatory neuron progression from Control to AD. F. Single-cell atlas-pie chart mapping the proposed chronological trajectory of excitatory neuron progression from Control to AD, with the pie chart representing the proportion of AD and Control in each subpopulation. G. Clustering-bubble chart demonstrating biological functions and pathways significantly activated by ExNeu_PRKN_VIRMA subpopulation genes. H. Pathway Map graph demonstrating the suppression of mitochondrial autophagy in ExNeu_PRKN_VIRMA.

### Increased abundance of PRKN and VIRMA double-positive subpopulations were also found in inhibitory neurons in AD brains

Similarly, five different cell subgroups were obtained by re-clustering the inhibitory neurons. Surprisingly, the subpopulation InNeu_PRKN_VIRMA, which is double positive for PRKN and VIRMA, was also found in inhibitory neurons, and again this subpopulation had the highest percentage in AD patients (Figure 3A). Subsequently, we explored the expression and co-expression of PRKN and VIRMA in the single-cell profiles of this subpopulation (Figure 3B). spatial transcriptome profiles of the InNeu_PRKN_VIRMA subpopulation showed that marker genes for inhibitory neurons *GAD1, PRKN* and *VIRMA* had higher and more widespread expression in AD compared to controls (Figure 3C). By cellular abundance analysis, we found that the abundance of ExNeu_PRKN_VIRMA subpopulation was likewise higher in AD (Figure 3D). The proposed chronological analysis showed that the cells of the InNeu_PRKN_VIRMA subpopulation of inhibitory neurons were partially in early development but mostly in late development during the development of AD (Figure 3E, F). Enrichment analysis showed that the InNeu_PRKN_VIRMA subpopulation’s were significantly involved in BP such as positive regulation of mitochondrial autophagy by mitochondrial depolarization, regulation of mitochondrial autophagy and mitochondrial to lysosomal transport as well as axon guidance, glutamatergic synapses, cell adhesion molecules, Rap1 signaling pathway and cAMP signaling pathway (Figure 3G). In addition, we can see that mitochondrial autophagy of InNeu_PRKN_VIRMA is inhibited in the pathway map (Figure 3H).

**Figure 3.**
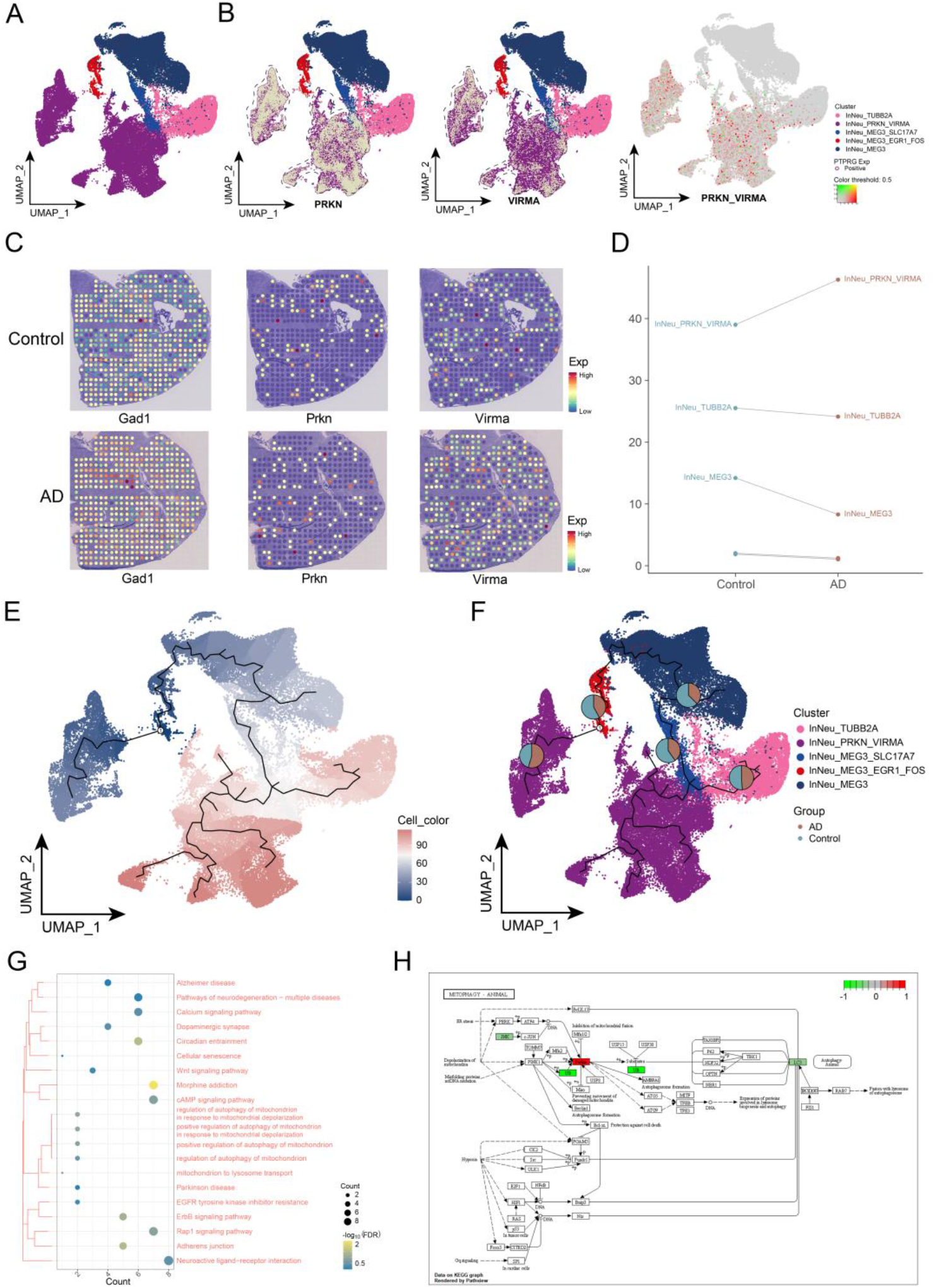
Role of inhibitory neuron subpopulations in the development of AD. A. Single-cell atlas mapping inhibitory neuron subpopulations. B. Single-cell atlas showing the expression of PRKN and VIRMA, co-expression of PRKN and VIRMA. C. ST atlas showing the expression of marker genes *GAD1, PRKN* and *VIRMA* in inhibitory neuron subpopulations. D. Dotted line graph showing proportion of inhibitory neuron subpopulations in AD-Control. E. Single-cell atlas mapping the proposed chronological values of inhibitory neuron progression from Control to AD. F. Single-cell atlas-pie chart mapping the proposed chronological trajectory of inhibitory neuron progression from Control to AD, with the pie chart representing the proportion of AD and Control in each subpopulation. G. Clustering-bubble chart demonstrating biological functions and pathways significantly activated by InNeu_PRKN_VIRMA subpopulation genes. H. Pathway Map graph demonstrating the suppression of mitochondrial autophagy in InNeu_PRKN_VIRMA.

### VIRMA-promoted m6A methylation of PRKN mediates mitochondrial damage depletion contributing to a major component of the neuronal death ecotone

Correlation analysis showed that the expression of VIRMA and PRKN were significantly positively correlated in excitatory and inhibitory neurons (Figure 4A, B). Not only that, the abundance of ExNeu_PRKN_VIRMA and InNeu_PRKN_VIRMA subpopulations were also significantly positively correlated in AD patient samples (Figure 4C), thus, VIRMA may have a reciprocal relationship with PRKN. Sequence prediction by catRAPID revealed a binding region of VIRMA protein to the mRNA of PRKN (Figure 4D). Subsequent molecular docking confirmed the possibility of binding of VIRMA protein to the mRNA of PRKN (Figure 4E), suggesting that VIRMA plays an important role in the neuronal death by promoting high PRKN expression mediating mitochondrial damage through m6A methylation.

**Figure 4.**
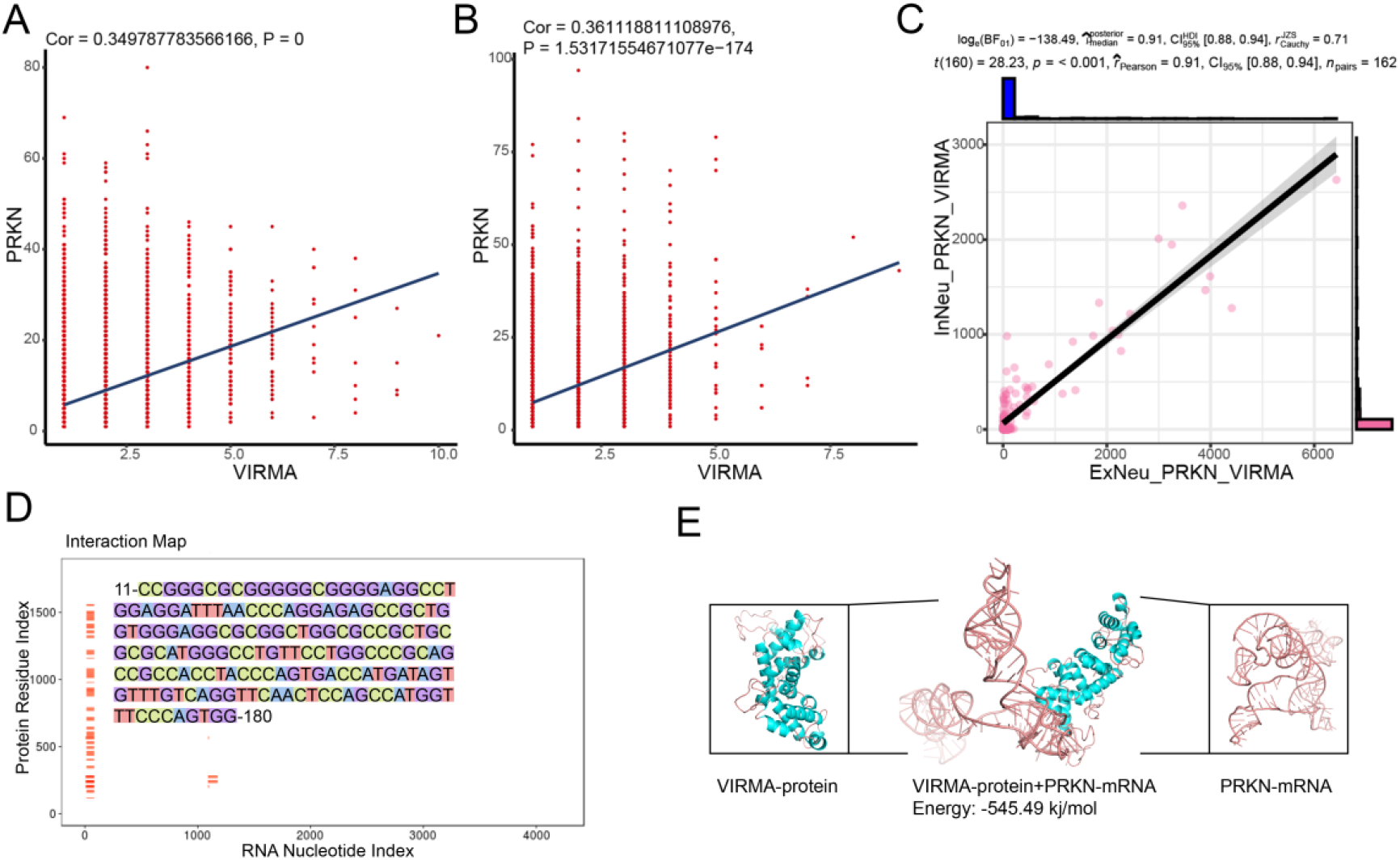
Validation of VIRMA and PRKN interactions. A. Correlation scatter plot demonstrating the correlation of VIRMA and PRKN expression in ExNeu. B. Correlation scatter plot demonstrating the correlation of VIRMA and PRKN expression in InNeu. C. Correlation scatter plot demonstrating the correlation of ExNeu_PRKN_VIRMA and InNeu_PRKN_ in patient samples. VIRMA abundance correlation. D. Heat map demonstrating the mRNA binding potential of VIRMA protein and PRKN. E. Molecular docking model demonstrating the binding sites of VIRMA protein and mRNA of PRKN.

### Mic_PTPRG is significantly increased with amyloid beta precursor protein deposition in AD brain

Twenty different cell subpopulations were obtained by re-clustering microglia (Figure 5A). Notably, the receptor protein tyrosine phosphatase (Ptprg-RPTPγ) encoded by PTPRG is predominantly expressed in the nervous system and has an important role in human diseases such as neuropsychiatric and behavioral disorders, central nervous system and inflammatory diseases. Therefore, we further explored the Mic_PTPRG subpopulation. As shown in Figure 5B, the expression of PTPRG was mapped in a single-cell atlas of microglia. ST mapping revealed that *PTPRG* expression was dysregulated in AD compared to control (Figure 5C). In addition, Mic_PTPRG subpopulation cells were also in higher abundance in AD (Figure 5D). Enrichment analysis revealed that the dysregulated genes in Mic_PTPRG subpopulation cells were significantly involved in mitochondrial DNA replication regulation and autophagic cell death regulatory processes (Figure 5E). The proposed chronological analysis showed that Mic_PTPRG subpopulation cells were at a late developmental stage in the progression of microglia to AD (Figure 5F,G).

**Figure 5.**
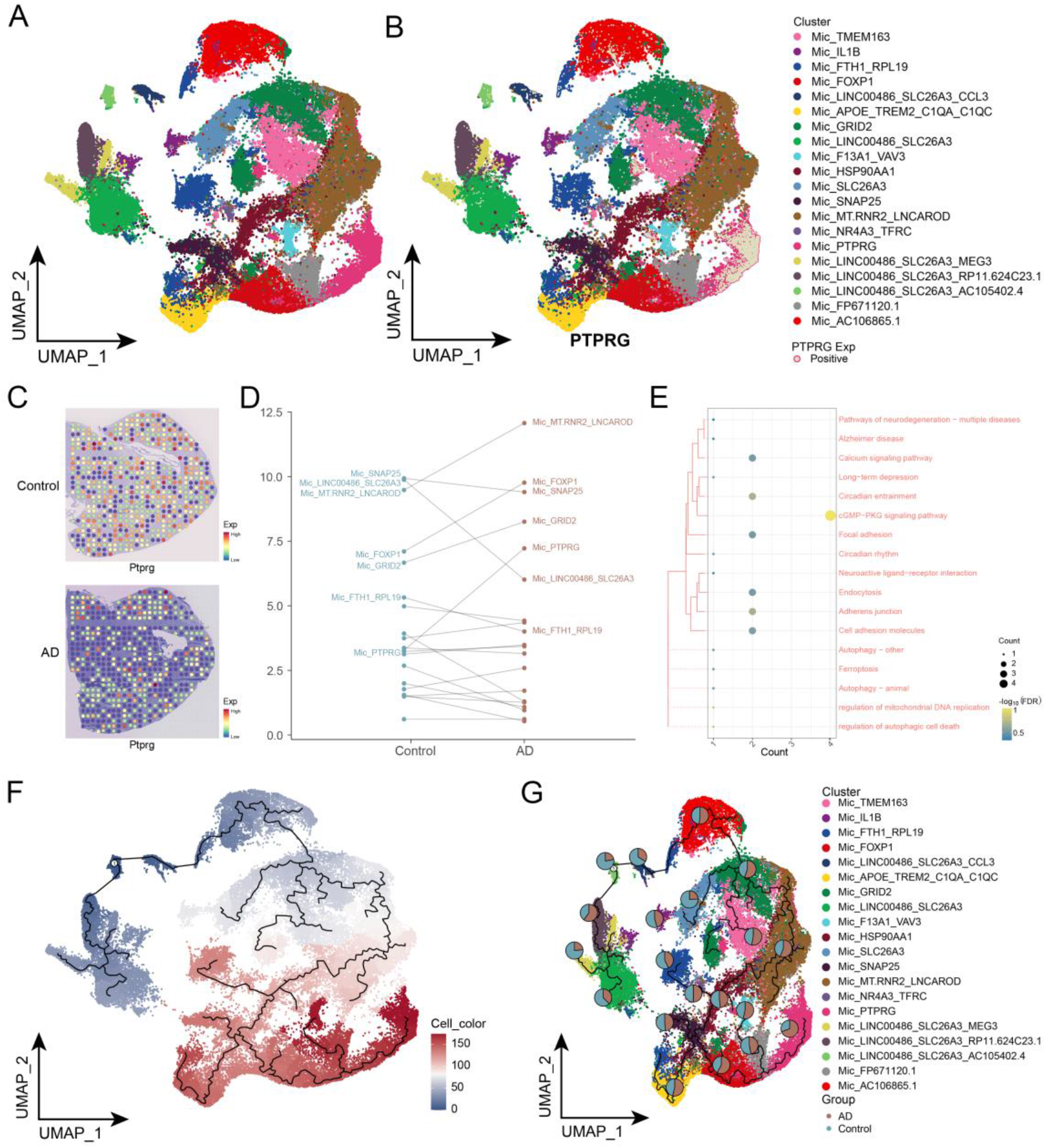
Role played by microglia subpopulations in AD development. A. Single-cell atlas mapping microglia subpopulations. B. Single-cell atlas mapping PTPRG expression showing microglia subpopulations. C. Vacancy mapping showing PTPRG expression in microglia subpopulations. D. Dotted line graph showing the proportion of microglia subpopulations in AD-Control. E. Clustered-bubble graph showing biological functions and pathways of significant gene activation in Mic_PTPRG subpopulation. F. Single-cell atlas mapping the proposed chronological values of microglia progression from Control to AD. G. Single-cell atlas-pie chart mapping the proposed chronological trajectory of microglia progression from Control to AD, with the pie chart representing the proportion of AD and Control in each subpopulation.

### Mic_PTPRG communicates intercellularly with neurons based on ligand-receptor interaction pairs

Compared to the control, the cellular abundance of the Mic_PTPRG subpopulation with the PRKN and VIRMA double-positive subpopulations InNeu_PRKN_VIRMA and ExNeu_PRKN_VIRMA were both significantly increased in AD and both were at a late stage of cell development in the progression to AD. Preliminary analysis revealed extensive communication between these subpopulations of cells (Figure 6A). Notably, both InNeu_PRKN_VIRMA and ExNeu_PRKN_VIRMA subpopulations were present in which CNTN4 was able to target the PTPRG of the Mic_PTPRG subpopulation (Figure 6B). Subsequently, the expression of CNTN4 and co-expression with PRKN and VIRMA, respectively, were mapped in the cell profiles of InNeu_PRKN_VIRMA and ExNeu_PRKN_VIRMA subpopulations. It can be observed that CNTN4 was widely expressed in InNeu_PRKN_VIRMA and ExNeu_PRKN_VIRMA subpopulations (Figure 6C, D). In addition, spatial transcriptome profiling showed higher and more extensive expression of *CNTN4* and *PTPRG* in AD compared to controls (Figure 6E). This suggests that the Mic_PTPRG subpopulation may communicate intercellularly with the InNeu_PRKN_VIRMA and ExNeu_PRKN_VIRMA subpopulations of neurons based on PTPRG-CNTN4.

**Figure 6.**
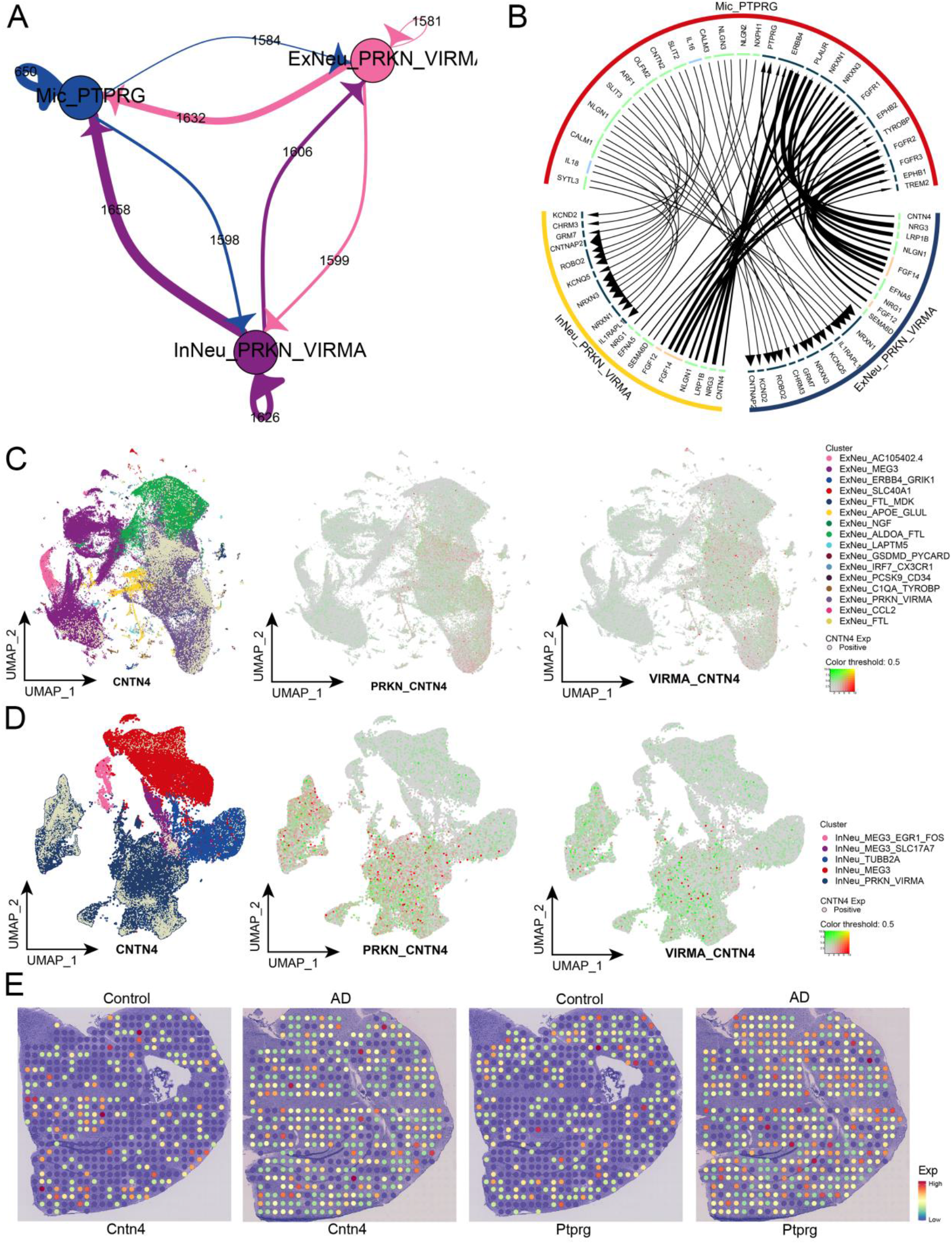
Verification of the existence of cellular communication between the Mic_PTPRG subpopulation and the PRKN and VIRMA double-positive subpopulations. A. Vesicular circular network diagram demonstrating the communication profile between microglia and VIRMA and PRKN double-positive neuronal cells (ExNeu_PRKN_VIRMA and InNeu_PRKN_VIRMA) in AD brain tissue. B. Reciprocal circular network plot demonstrating the communication events between Mic_PTPRG and VIRMA and PRKN dual positive neuronal cells (i.e. ExNeu_PRKN_VIRMA and InNeu_PRKN_VIRMA) in AD brain tissue. C. Single-cell atlas demonstrating CNTN4 expression in ExNeu_PRKN_VIRMA and CNTN4 and PRKN_ VIRMA co-expression. D. Single-cell profiles demonstrating CNTN4 expression in InNeu_PRKN_VIRMA and co-expression of CNTN4 and PRKN_VIRMA. E. ST atlas demonstrating *CNTN4* and *PTPRG* expression.

### Ectopic expression of microglia-derived PTPRG induces PRKN and VIRMA double-positive neuronal production

Similarly, we observed that PTPRG was widely expressed in the InNeu_PRKN_VIRMA and ExNeu_PRKN_VIRMA subpopulations and co-expressed with PRKN and VIRMA (Figure 7A, B). Sequence prediction by catRAPID revealed a binding region of PTPRG protein to the mRNA of VIRMA (Figure 7C), and molecular docking confirmed the possibility of binding of PTPRG protein to the mRNA of VIRMA (Figure 7D). It is thus speculated that there may be ectopic expression of microglia-derived PTPRG, which induces the production of PRKN and VIRMA double-positive neurons.

**Figure 7.**
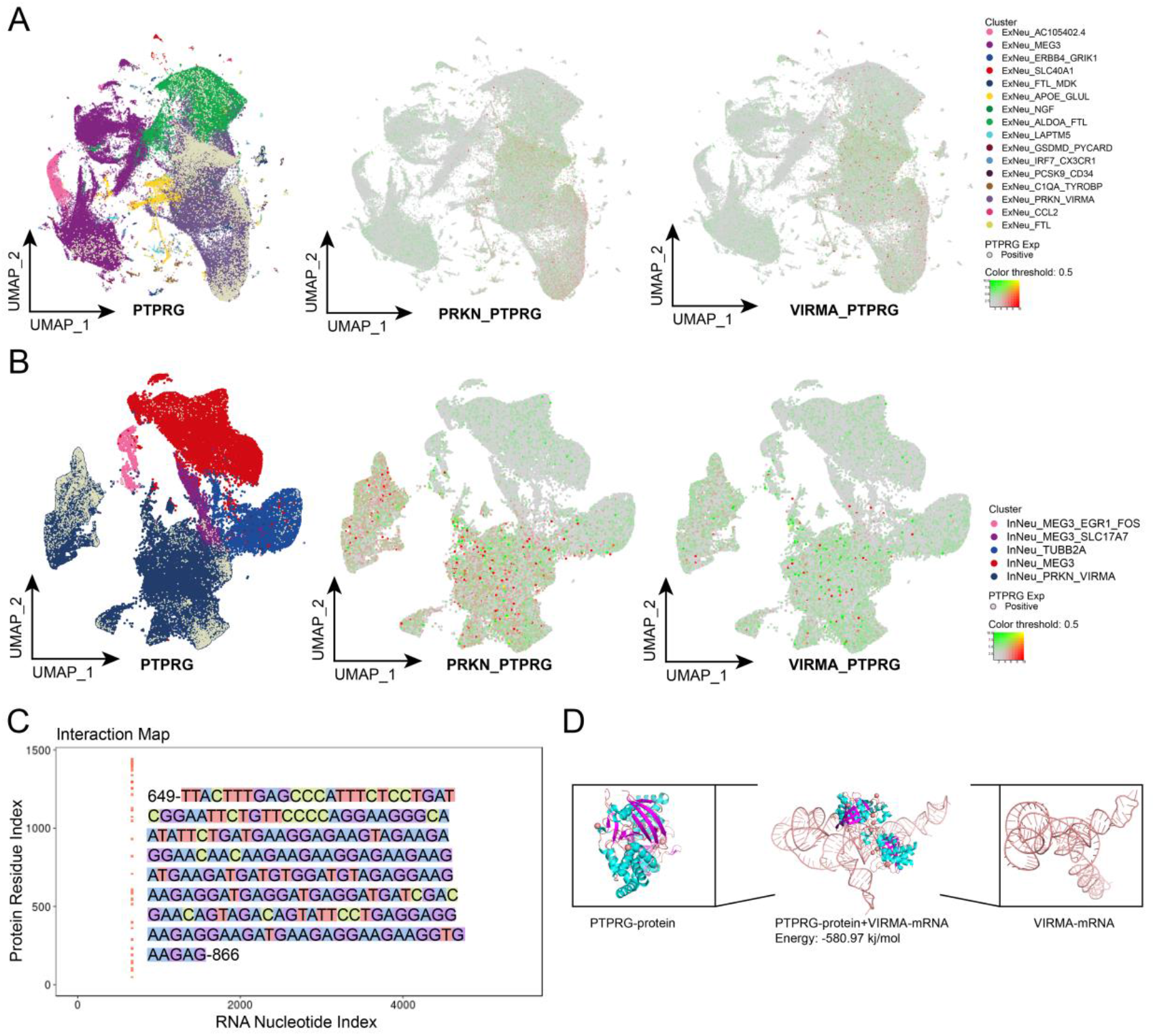
Validation of PTPRG binding to VIRMA ligand-receptor. A. Single-cell mapping demonstrating the expression of PTPRG in ExNeu_PRKN_VIRMA and co-expression of PTPRG and PRKN_VIRMA. B. Single-cell mapping demonstrating the expression of PTPRG in InNeu_PRKN_VIRMA and co-expression of PTPRG and PRKN_VIRMA. C. Heat map demonstrating the mRNA binding potential of PTPRG protein and VIRMA. D. Molecular docking model demonstrating the binding sites of PTPRG protein and mRNA of VIRMA.

## Discussion

Using the single-cell landscape constructed in this study, we found a significant dysregulation of mitochondrial gene expression in both excitatory and inhibitory neurons in AD compared to controls, and possible neuronal mitochondrial damage depletion-mediated cell death. A series of neuronal subpopulations were subsequently identified and validated in spatial histology. Among them, the ExNeu_PRKN_VIRMA subpopulation had the highest proportion in AD patients and progressively increased in abundance over the progression of AD. Intriguingly, a double positive subpopulation of PRKN and VIRMA was also found in inhibitory neurons, and its abundance also increased progressively during the progression of AD. Various degrees of dysregulated mitochondrial gene expression were found in both ExNeu_PRKN_VIRMA and InNeu_PRKN_VIRMA neuronal subpopulations. Through further analysis, these neuronal subpopulations were found to be the main constituents contributing to the ecology of mitochondrial damage depletion in AD neurons with certain mitochondrial damage depletion as well as energy and nutrient metabolic imbalance that induces neuronal death.

Mitochondria are organelles essential for synaptic function, and mitochondrial dysfunction and synaptic failure are also characteristic early manifestations of pathology in the brain of AD patients (26). There is growing evidence that mitochondrial disorders are key factors contributing to synaptic failure and degeneration in AD (27, 28), and proteomic studies of synaptic vesicles in AD brain tissue have also highlighted mitochondrial dysfunction at synapses (29, 30). Significant changes in mitochondrial dynamics and organization, such as being elongated, enlarged and over fusion, also occur in different models of cellular senescence (31). Furthermore, it was shown that PRKN (parkin RBR E3 ubiquitin protein ligase) plays a key role in mitochondrial autophagy regulation and neuroprotection (32) and its overexpression inhibits extracellular mitochondrial release under basal and stress conditions (33). In the early and middle stages of AD, Parkin levels increased significantly and the recruitment to depolarized mitochondria increased, promoting mitochondrial autophagy and clearance of damaged mitochondria, but with disease progression, Parkin levels gradually decreased and eventually depleted, which caused a large accumulation of defective mitochondria and eventually affected neuronal function, leading to their death (34). PINK1 (PTEN-inducible kinase 1) recruits PRKN to depolarized mitochondria that ubiquitinates multiple target proteins on the outer mitochondrial membrane and subsequently promotes autophagic degradation of mitochondria (35, 36). In contrast, disruption of PINK1/Parkin signaling leads to impaired mitochondrial function and ultimately to neurodegenerative and neuroinflammatory processes (37). This suggests that PINK1-recruited PRKN can activate mitochondrial autophagy and mitotic clearance of damaged mitochondria and protect neurons.

However, in our study, none of these neuronal subpopulations expresses PINK1 or activates mitochondrial autophagy. It is possible that the progression of AD is accompanied by defective mitochondrial autophagy and consequent progressive depletion of PRKN, which in turn exacerbates the ecology of neuronal death. However, the mechanism of PRKN depletion remains unexplored due to the uncertainty of AD progression and changes in molecular abundance. As we captured a large subpopulation of PRKN transcript-positive neurons in AD patients, it provides us with the conditions to observe and explore the molecular mechanism of PRKN depletion at single-cell resolution. In the PRKN transcript-positive neuronal subpopulations, PRKN and the m6A methyltransferase VIRMA were co-expressed in the ExNeu and InNeu subpopulations, suggesting a potential association between the two. Among them, VIRMA is a related gene encoding a Vir-like m6A methyltransferase. In contrast, n6-methyladenosine (m6A) is the most common chemical epigenetic modification in mRNA post-transcriptional modifications, including methylation, demethylation and recognition (38). In the healthy brain, m6A maintains a variety of developmental and physiological processes such as neurogenesis, axonal growth, synaptic plasticity, circadian rhythms, cognitive function, and stress response (39). Aberrant expression of m6A-related proteins may occur in the nervous system, thereby affecting neurogenesis, brain volume, learning memory, memory formation, and consolidation (40). By constructing a molecular docking model, we identified the binding potential of VIRMA to PRKN. This suggests that VIRMA may promote the degradation of PRKN transcripts by increasing their m6A methylation modification levels to inhibit their translation.

Multiple studies have collectively shown that microglia are critical for neuronal homeostasis and health (41). In the adult brain, microglia have multiple functions, including monitoring changes in neuronal activity, regulating learning and memory, and acting as local phagocytosis and injury sensors in the brain parenchyma (42-48). Among them, mostly microglia-neuron interactions are mediated by cell-cell signaling pathways, including purinergic signaling pathways, cytokines, neurotransmitters and neuropeptides (49). Previous studies have shown that accumulation of pathological Aβ or Tau at the synapse can upregulate C1q in microglia and promote complement activation and subsequent microglia phagocytosis at the synapse (50-53). Blocking activation of the classical complement cascade pathway in mouse models of AD with genetic or antibody-based approaches has been shown to protect synapses from synaptic loss, dysfunction, and memory loss (54), suggesting that the microglia-synaptic pathway may be a potential therapeutic target.

Through intercellular communication analysis, we identified that Mic_PTPRG, a subpopulation of microglia also progressively increased in AD progression, can anchor target and communicate with PRKN and VIRMA dual positive neuronal subpopulations based on ligand-receptor interaction pairs, which in turn promotes ectopic expression of PTPRG in neurons, while aberrantly expressed PTPRG enhances the RNA of VIRMA stability. Notably, the receptor protein tyrosine phosphatase (Ptprg-RPTPγ) encoded by PTPRG is predominantly expressed in the nervous system and has an important role in human diseases such as neuropsychiatric and behavioral disorders, central nervous system and inflammatory diseases (55), and is a potential risk gene for late-onset dementia (56). PTPRG mediates cell adhesion and signaling during development (57), which is usually upregulated during neuroinflammation (58) while microglia are specialized phagocytes that reside in tissues for long periods of time and investigate and rapidly respond to changes in their microenvironment (59). It is thus hypothesized that the microglia subpopulation Mic_PTPRG may arise due to microglial role in phagocytosis and removal of over-deposited APP from the patient’s brain.

Given certain limitations, our results should be interpreted with caution. Given our results were obtained exclusively by bioinformatics analysis, they should be validated and extended in a functional biochemical study.

## Conclusion

Our study shows that excessive deposition of amyloid β precursor protein can induce PTPRG-positive microglia subpopulations to produce and anchor target neurons via intercellular ligand-receptor interactions communication, causing ectopic expression of PTPRG in neurons. PTPRG in neurons can bind and upregulate the expression of m6A methyltransferase VIRMA, which in turn increases the level of m6A modification of PRKN transcripts and decreases their RNA stability and translational activity. Expression of repressed PRKN causes a clear block in neuronal cell injury depleting mitochondria and builds up, which in turn reprograms neuronal energy and nutrient metabolic pathways and mediates neuronal death under AD progression.

## Data Availability

All data produced in the present study are available upon reasonable request to the authors.

## Data availability statement

The datasets supporting the conclusions of this article are available in the Gene Expression Omnibus (www.ncbi.nlm.nih.gov/geo).

## Ethics Approval and Informed Consent

The Ethics Committee from the Second Affiliated Hospital of Guangxi Medical University waived the requirements for ethics approval because our analyses were based entirely on publicly available data from studies that had been approved by the relevant ethics committees at the research institutions involved.

## Acknowledgement

This study was supported by the National Natural Science Foundation of China (82060210), the Nanning Excellent Young Scientist Program and the Guangxi Beibu Gulf Economic Zone Major Talent Program (RC20190103).

